# Evaluation of Large Language Model Performance on the Biomedical Language Understanding and Reasoning Benchmark: Comparative Study

**DOI:** 10.1101/2024.05.17.24307411

**Authors:** Hui Feng, Francesco Ronzano, Jude LaFleur, Matthew Garber, Rodrigo de Oliveira, Kathryn Rough, Katharine Roth, Jay Nanavati, Khaldoun Zine El Abidine, Christina Mack

**Affiliations:** Real world solutions, IQVIA

**Keywords:** artificial intelligence, large language model, large language models, LLM, natural language processing, NLP, ChatGPT

## Abstract

**Background:** The availability of increasingly powerful large language models (LLMs) has attracted substantial interest in their potential for interpreting and generating human-like text for biomedical and clinical applications. However, there are often demands for high accuracy, concerns about balancing generalizability and domain-specificity, and questions about prompting robustness when considering the adoption of LLMs for specific use cases. There also is a lack of a framework or method to help choose which LLMs (or prompting strategies) should be adopted for specific biomedical or clinical tasks.

**Objective:** To address the speculations on applying LLMs for biomedical applications, this study aims to 1) propose a framework to comprehensively evaluate and compare the performance of a range of LLMs and prompting techniques on a suite of biomedical natural language processing (NLP) tasks; 2) use the framework to benchmark several general-purpose LLMs and biomedical domain-specific LLMs.

**Methods:** We evaluated and compared six general-purpose LLMs (GPT-4, GPT-3.5-Turbo, Flan-T5-XXL, Llama-3-8B-Instruct, Yi-1.5-34B-Chat, and Zephyr-7B-Beta) and three healthcare-specific LLMs (Medicine-Llama3-8B, Meditron-7B, and MedLLaMA-13B) on a set of 13 datasets – referred to as the Biomedical Language Understanding and Reasoning Benchmark (BLURB) – covering six commonly needed medical natural language processing tasks: named entity recognition (NER); relation extraction (RE); population, interventions, comparators, and outcomes (PICO); sentence similarity (SS); document classification (Class.); and question-answering (QA). All models were evaluated without further training or fine-tuning. Model performance was assessed according to a range of prompting strategies (formalized as a systematic, reusable prompting framework) and relied on the standard, task-specific evaluation metrics defined by BLURB.

**Results:** Across all tasks, GPT-4 outperformed other LLMs, achieving a score of 64.6 on the benchmark, though other models, such as Flan-T5-XXL and Llama-3-8B-Instruct, demonstrated competitive performance on multiple tasks. We found that general-purpose models achieved better overall scores than domain-specific models, sometimes by significant margins. We observed a substantial impact of strategically editing the prompt describing the task and a consistent improvement in performance when including examples semantically similar to the input text. Additionally, the most performant prompts for nearly half the models outperformed the previously reported best results for the PubMedQA dataset from the BLURB leaderboard.

**Conclusions:** These results provide evidence of the potential LLMs may have for biomedical applications and highlight the importance of robust evaluation before adopting LLMs for any specific use cases. Notably, performant open-source LLMs such as Llama-3-8B-Instruct and Flan-T5-XXL show promise for use cases where trustworthiness and data confidentiality are concerns, as these models can be hosted locally, offering better security, transparency, and explainability. Continuing to explore how these emerging technologies can be adapted for the healthcare setting, paired with human expertise, and enhanced through quality control measures will be important research to allow responsible innovation with LLMs in the biomedical area.

## Introduction

Recent advances in large language models (LLMs) have generated substantial interest in using them to perform natural language processing (NLP) tasks in the medical domain, including writing clinical notes, summarizing scientific literature, and reasoning about public health topics [1].

There are also serious concerns about the safety, ethics, and trustworthiness of LLM outputs [2], with evidence of plausible – but factually incorrect – hallucinations of medical information [3], perpetuation of racial bias [4], and omission of important information from LLM summaries [5]. Calls to rigorously evaluate LLM performance when performing specific medical tasks [6] emphasize the need to ensure adequate information is available for evidence-based decision-making when these technologies will impact patient care and treatment.

In addition to the need for robust evaluation, there are open questions about how to prompt models to ensure quality outputs for specific biomedical tasks and to what extent domain-specific training of models is beneficial. Significant performance increases have been observed when the instructions for the task are accompanied by several examples (few-shot learning), compared to only providing instructions (zero-shot learning) [7].

Example selection strategies to populate few-shot prompts can also meaningfully impact performance, specifically by selecting semantically similar examples using sentence-level embeddings created by models such as BERT (Bidirectional Encoder Representations from Transformers) [8] [9]. Another strategy for improving LLM performance is fine-tuning with data from the biomedical domain. While this approach requires substantial compute resources, specialized technical expertise, and a relatively large amount of high-quality training data, it can be worthwhile when it leads to increased quality and trustworthiness of the LLM’s output. Furthermore, this type of domain adaptation can enable smaller models to match or surpass the performance of larger general-purpose ones on domain-specific tasks. For example, Wu et al trained MedLLaMA by adapting the LLaMA model on a corpus of biomedical papers and medical textbooks, and then created PMC-LLaMA by further fine-tuning it on an instruction dataset comprised of medical NLP tasks [10]. Both models outperformed ChatGPT on multiple medical question-answering benchmarks, despite having significantly fewer parameters.

To help answer these questions, we conducted an empirical evaluation to assess the ability of nine LLMs – six general-purpose and three domain-specific (ie, trained on biomedical texts) – to perform biomedical NLP tasks, all of which are included in the Biomedical Language Understanding and Reasoning Benchmark (BLURB) [11]. In addition to benchmarking, we investigated how the quality of LLM responses can be improved through prompting strategies.

## Methods

### BLURB Benchmarking Datasets and Healthcare-Based Tasks

BLURB encompasses 13 publicly available biomedical NLP datasets used to evaluate the following six common biomedical NLP tasks:

- **Named entity recognition (NER)** refers to the extraction of specified categories of information from text – such as identifying all mentions of medications, diseases, or cell types. Associated datasets are all based on PubMed abstracts with specific annotated mentions: *BC5-chem:* drugs and chemical compounds; *BC5-disease:* diseases; *NCBI-disease:* diseases; *BC2GM:* genes; *JNLPBA:* molecular biology concepts (ie, protein, DNA, RNA, cell line, cell type).
- **Population, interventions, comparators, and outcomes (PICO)** identifies each of the categories from the PICO research framework from an abstract [12]. The PICO dataset, *EMB-PICO,* is a collection of clinical trial abstracts with annotated mentions of these key elements of study design.
- **Relation extraction (RE)** is the task of classifying the relationship between pairs of entities in a text. There are three such datasets in BLURB: *ChemProt* is a collection of PubMed abstracts with pairs of chemical and protein entities classified into several different relationships; *DDI* is a collection of texts from PubMed abstracts and DrugBank with pairs of drugs annotated with different types of drug-drug interactions; *GAD* is a collection of sentences from PubMed abstracts classified as to whether they contain a gene-disease relationship.
- **Sentence similarity (SS)** is the task of determining the similarity of the meaning of two sentences. The sentence similarity dataset, *BIOSSES,* is a collection of sentence pairs that have been rated on a scale of 0 (no relation) to 4 (equivalent meanings) by subject matter experts, which the model must correctly estimate.
- **Document classification (Class.)** is the task of categorizing a document. The document classification dataset, *HoC,* is a collection of PubMed abstracts that have been classified according to whether they discuss specific cancer hallmarks.
- **Question answering (QA)** is the task of correctly answering free-text questions. The two question answering datasets are both based on PubMed abstracts and annotated with question-answer pairs based on the provided text: *PubMedQA* captures whether the text contains the answer to a research question (yes/no/maybe); *BioASQ* annotates whether an extract is the correct answer to a research question (yes/no).

Each dataset has clearly defined ground truth labels and was partitioned into training, validation, and test sets as part of the BLURB benchmark; all evaluations reported here were performed on the standard BLURB test set using the provided labels and pre-defined evaluation metrics. Further detail on BLURB tasks is available [11] and information is summarized in (Table 1).

**Table 1.** Overview of tasks, metrics, and datasets for the Biomedical Language Understanding and Reasoning Benchmark (BLURB).

| Task | Metric | Datasets |
| --- | --- | --- |
| Named-Entity Recognition (NER) | F1 entity-level:<br>F1 score calculated at named entity level. | BioCreative II Gene Mention Recognition (*BC2GM*) |
|  |  | BC5CDR Drug/Chemical (*BC5-chem*) |
|  |  | BC5CDR Disease (*BC5-disease*) |
|  |  | *JNLPBA*: chemical recognition |
|  |  | *NCBI Disease*: disease recognition |
| Populations, Interventions, Comparators, and Outcomes (PICO) | Macro F1 word-level:<br>Unweighted average of F1 score across the four classes. | *EBM-PICO*: extract PICO from medical abstracts describing clinical trials |
| Relation Extraction (RE) | Micro F1:<br>Global average of F1 score for all mentions of relationships. | *ChemProt*: chemical-protein interactions |
|  |  | Drug-Disease Interaction (*DDI*) |
|  |  | Gene-Disease Associations (*GAD*) |
| Sentence Similarity (SS) | Pearson coefficient:<br>The covariance of the two variables divided by the product of their standard deviations. | *BIOSSES*: biomedical sentence similarity |
| Document Classification (Class.) | Average micro F1:<br>Unweighted average of micro F1 score for all classes. | Hallmarks of Cancer (*HoC*) |
| Question Answering (QA) | Accuracy:<br>Number of correct predictions divided by total number of predictions. | *BioASQ*: biomedical QA |
|  |  | *PubMedQA*: biomedical QA based on PubMed abstracts |

### Models

Nine LLMs were compared: six general-purpose LLMs (Azure-based GPT-3.5-Turbo and GPT-4 [13], Flan-T5-XXL [14], Llama-3-8B-Instruct [15], Yi-1.5-34B-Chat [16], and Zephyr-7B-Beta [17]) and three adapted for the biomedical domain (Medicine-Llama3-8B [18] [19], Meditron-7B [20], and MedLLaMA-13B [10]). GPT-3.5-Turbo and GPT-4 are closed-source commercial models, while the remainder of the models are open-source. Among the domain-specific models, Meditron-7B and MedLLaMA-13B were created by further pre-training a base model on a variety of clinical and medical text data sources; Medicine-Llama3-8B was created by training on a corpus of PubMed abstracts enhanced with instruction pre-training, a process in which the original corpus is augmented with automatically generated instruction-response pairs [18]. Additional details for each of the nine LLMs can be found in (Table 2). None of the models underwent further training or fine-tuning for this evaluation.

**Table 2.** Model details for each of the nine LLMs evaluated.

| Model | # Parameters | Architecture | Domain | Instruction-tuned | Open-source |
| --- | --- | --- | --- | --- | --- |
| Flan-T5-XXL | 11B | Encoder-decoder | General-purpose | Yes | Yes |
| GPT-3.5-Turbo | Undisclosed | Decoder-only | General-purpose | Yes | No |
| GPT-4 | Undisclosed | Decoder-only | General-purpose | Yes | No |
| Llama-3-8B-Instruct | 8B | Decoder-only | General-purpose | Yes | Yes |
| MedLLaMA-13B | 13B | Decoder-only | Biomedical | No | Yes |
| Medicine-Llama3-8B | 8B | Decoder-only | Biomedical | Yes<sup>a</sup> | Yes |
| Meditron-7B | 7B | Decoder-only | Biomedical | No | Yes |
| Yi-1.5-34B-Chat | 34B | Decoder-only | General-purpose | Yes | Yes |
| Zephyr-7B-Beta | 7B | Decoder-only | General-purpose | Yes | Yes |

<sup>a</sup>Trained with instruction pre-training, rather than standard supervised fine-tuning.

### Prompting Strategies

A series of experiments were conducted to systematically evaluate prompting strategies based on the structure of the prompt and example selection. Each of the nine LLMs were provided with prompts constructed from four high-level templates (based on studies from previous work [21] [22]) across all datasets to assess the impact of distinct combinations of three key variables:

- **Verbosity** (*short versus long*): Short prompts contain concise instructions to the model. Long prompts provide additional details (eg, a description of each label) that could be potentially useful to the model.
- **Number of examples** (*zero-shot versus few-shot)*: Zero-shot prompts provide instructions to the model with no examples. Few-shot prompts provided the model with three examples of inputs and the corresponding correct responses.
- **Example selection strategy** (*semantically similar versus random*): Previous studies suggest that when prompting an LLM to analyze a specific text excerpt, quality of the LLM response can be improved by including examples in the prompt that are semantically similar to the text excerpt to be analyzed [8] [9]. Based on these studies, we implemented two approaches for example selection from the training set: semantically similar selection scheme and random selection scheme. Under the first scheme, we compute the sentence-level embeddings (or entity-level embeddings, for NER) using SapBERT [23] for training examples and select the three most similar to input text. We provide these examples along with their correct answers to the LLM as part of the prompt. For NER datasets, among the three semantically similar examples, we included two positive examples (ie, ground truth contains target entities) and one negative example (ie, ground truth contains no entities); for all other datasets, we simply used the three most similar examples. Under the random selection scheme, three examples were randomly selected.

For all the few-shot experiments for the EBM-PICO dataset and for the few-shot experiments for PubMedQA with Meditron-7B, MedLLaMA-13B, and Flan-T5-XXL, we encountered the issue of prompt length exceeding maximum context size of some models, and therefore did not conduct the experiments.

For most tasks, each model was prompted only once per example per prompting strategy. However, for EBM-PICO we prompted the model separately for each category (population, interventions, and comparators) and combined the responses before scoring. Additionally, for ChemProt and DDI we prompted each model separately for each pair of entities in the sentence.

The precise prompt texts used in our experiments are provided in (Multimedia Appendix 1).

### Evaluation Methodology

For each LLM and each dataset, six experiments were run varying the length of the prompt, the number of examples, and the example selection strategy. Similar prompts were used to interact with all LLMs, with different prompts being used for chat models; full prompts and the models they correspond to are available in (Multimedia Appendix 1). An overview of the evaluation strategy is provided in (Figure 1).

**Figure 1.**
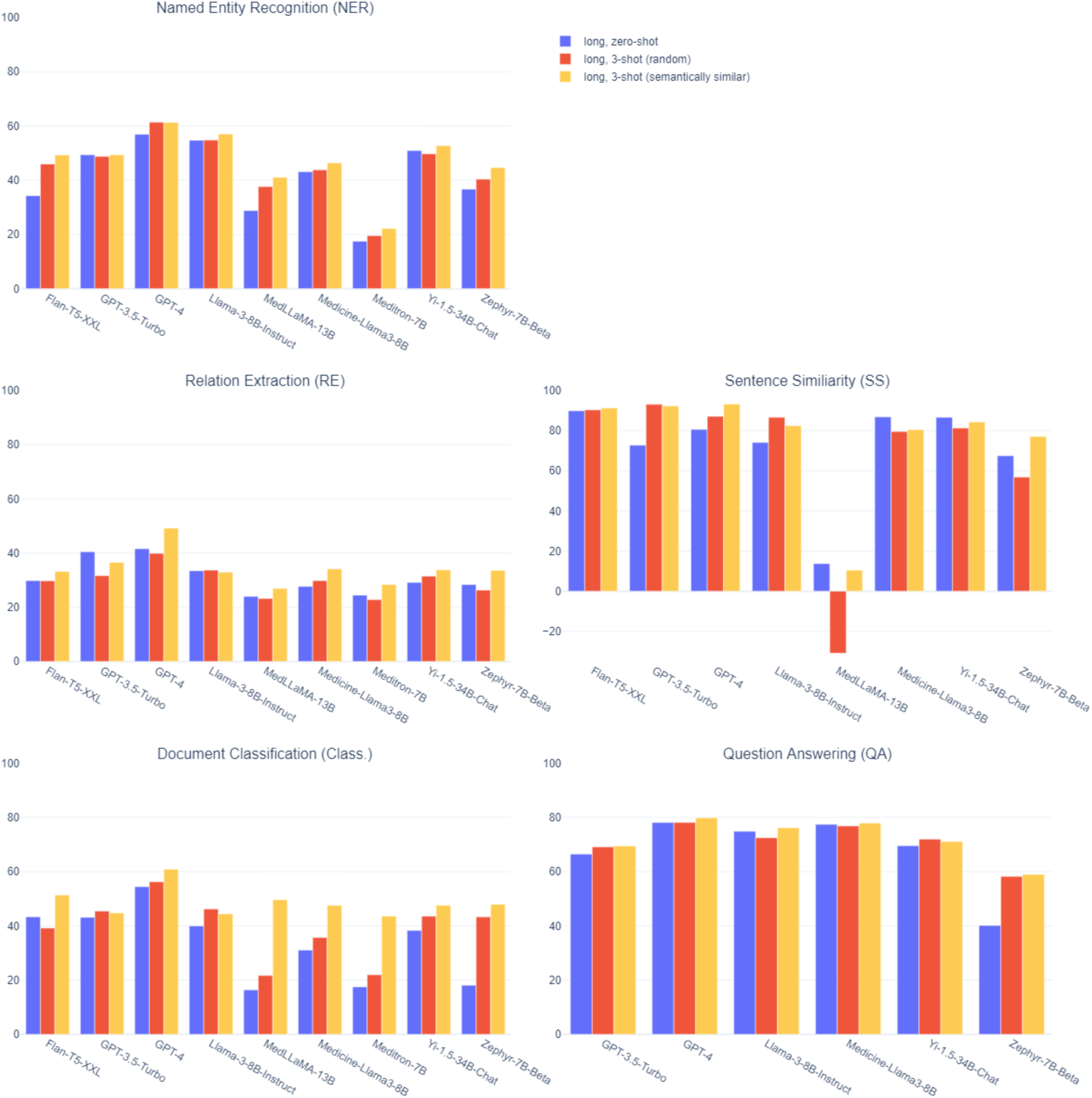
An illustration of the methodology used for this evaluation of the Biomedical Language Understanding and Reasoning Benchmark (BLURB) tasks.

To enforce standardization and repeatability of evaluations, raw responses from LLMs were formatted according to a standard JSON schema for each BLURB dataset. A detailed explanation of this process can be found in (Multimedia Appendix 2).

For all experiments performed, the entire test set was used to evaluate performance for a given dataset using the standard, task-specific metrics from the BLURB benchmark (see (Table 1)). For NER datasets, the F1 score (the harmonic mean of sensitivity and positive predictive value [ie, recall and precision]) was calculated. For the PICO task, we used a generalization of this metric, the macro F1 score (an unweighted mean of the F1 score for each of the three information categories). The micro F1 score – a weighted mean of category-specific F1 scores – was used for relation extraction and document classification. Pearson correlation coefficients were calculated to evaluate performance on the sentence similarity task. For the question answering task, accuracy – the overall proportion of correct answers – was used.

All analyses were performed using Python (3.9 and 3.10) packages, with scikit-learn and scipy for computing the metrics. The version of GPT models used was gpt-35-turbo, 0613 and gpt-4, 0613. Please reach out to the authors for scripts used to perform the evaluations.

## Results

(Multimedia Appendix 3) characterizes the performance of the nine LLMs against the test set of each BLURB dataset. For NER, relation extraction, and PICO datasets, the number of examples contained in each dataset varies from hundreds to thousands or tens of thousands. Document classification and question-answering datasets contained hundreds of examples. Sentence similarity contained twenty examples.

For all models and datasets, performance varied widely based on the prompting strategy used (Multimedia Appendix 3). For example, for GPT-4, the differences between the highest and lowest scores were typically less than 5 points for NER and PICO datasets but were much larger for the three relation extraction datasets (ranging from 9.55 to 26.34 points). For the sentence similarity and document classification datasets, the difference was about 12 points, and was less than 8 points for question-answering. However, this pattern was not generalizable across models; for example, the differences between highest and lowest scores for Flan-T5-XXL were from 10 to 20 points for the NER datasets and less than 8 points for the relation extraction datasets.

Although there was no single best prompting strategy across models and tasks, some clear patterns emerged. For seven of nine models, the long, few-shot prompting strategy was most commonly the most performant across all datasets. Additionally, depending on the dataset or task, the optimal prompting strategy appeared to make little difference in models’ performance. For example, there was little variation between the best and worst prompt for BioASQ, with the difference being just above 6 points when averaging over models. This contrasts with datasets such as NCBI-disease, for which this difference was nearly 16.5 points.

(Table 3) summarizes the average best performance of each model by task. We consider each model, prompt, and dataset as one combination and report the average per-model score across datasets of the same task. We also report the BLURB score for each model, which is the mean performance across tasks. Overall, GPT-4 showed the highest average best performance for all tasks. This was followed by Llama-3-8B-Instruct and GPT-3.5-Turbo, which had nearly identical BLURB scores. The highest performing biomedical LLM was Medicine-Llama3-8B, which scored about 5 points lower than its general-purpose equivalent. The two other biomedical models had the lowest BLURB scores, trailing the next-best LLM (Zephyr-7B-Beta) by over 10 points, and consistently performing worse than other models across tasks.

**Table 3.** Normalized mean of task-specific scores for the best-performing prompt for each large language model (LLM) for the Biomedical Language Understanding and Reasoning Benchmark (BLURB).

| Task<sup>a</sup> | Flan-T5-XXL | GPT-3.5-Turbo | GPT-4 | Llama-3-8B-Instruct | MedLLaMA-13B | Medicine-Llama3-8B | Meditron-7B | Yi-1.5-34B-Chat | Zephyr-7B-Beta |
| --- | --- | --- | --- | --- | --- | --- | --- | --- | --- |
| NER | 52.25 | 52.80 | 63.00 | 58.22 | 42.63 | 46.94 | 23.83 | 54.27 | 44.73 |
| PICO | 28.42 | 23.78 | 33.49 | 28.28 | 10.80 | 18.64 | 4.37 | 25.92 | 14.32 |
| RE | 33.21 | 41.33 | 50.54 | 34.32 | 26.93 | 34.17 | 29.15 | 34.60 | 33.91 |
| SS | 91.20 | 93.02 | 93.18 | 86.57 | 28.08 | 87.00 | 24.64 | 87.64 | 77.04 |
| Class. | 51.36 | 57.57 | 66.81 | 54.24 | 49.65 | 47.59 | 48.82 | 50.34 | 51.74 |
| QA | 70.55 | 72.42 | 80.56 | 79.33 | 62.35 | 77.90 | 57.73 | 74.69 | 61.85 |
| BLURB Score | 54.50 | 56.82 | 64.60 | 56.83 | 36.74 | 52.04 | 31.42 | 54.58 | 47.26 |

<sup>a</sup>NER: Named-Entity Recognition; PICO: Populations, Interventions, Comparators, and Outcomes; RE: Relation Extraction; SS: Sentence Similarity; Class.: Document Classification; QA: Question Answering

### Prompting Strategies: Impact of Short Versus Long Instructions

(Figure 2) compares the mean per-task performance of long versus short instructions by model when no examples are included in the prompt. There is no clear best strategy across tasks and models. In some cases, providing longer prompts dramatically improved the performance of a model. For example, for the sentence similarity task, considering the zero-shot scenarios, long instructions improved the MedLLaMA-13B model performance by 16.36 points and the Zephyr-7B-Beta model by 52.27 points; yet for the documentation classification task, more verbose prompts improved MedLLaMA-13B performance by 15.63 points but decreased the performance of Zephyr-7B-Beta by 26.00 points. For other models, like Flan-T5-XXL and GPT-4, more verbose instructions tended to decrease performance, as observed in four out of six tasks. For the relation extraction task, all models saw increased performance from the longer instructions, though the magnitude of the improvement varied (Figure 2). Using longer prompts negatively affected over half of the models for the PICO and document classification tasks. For all other tasks, changes in performance were mixed across models.

**Figure 2.**
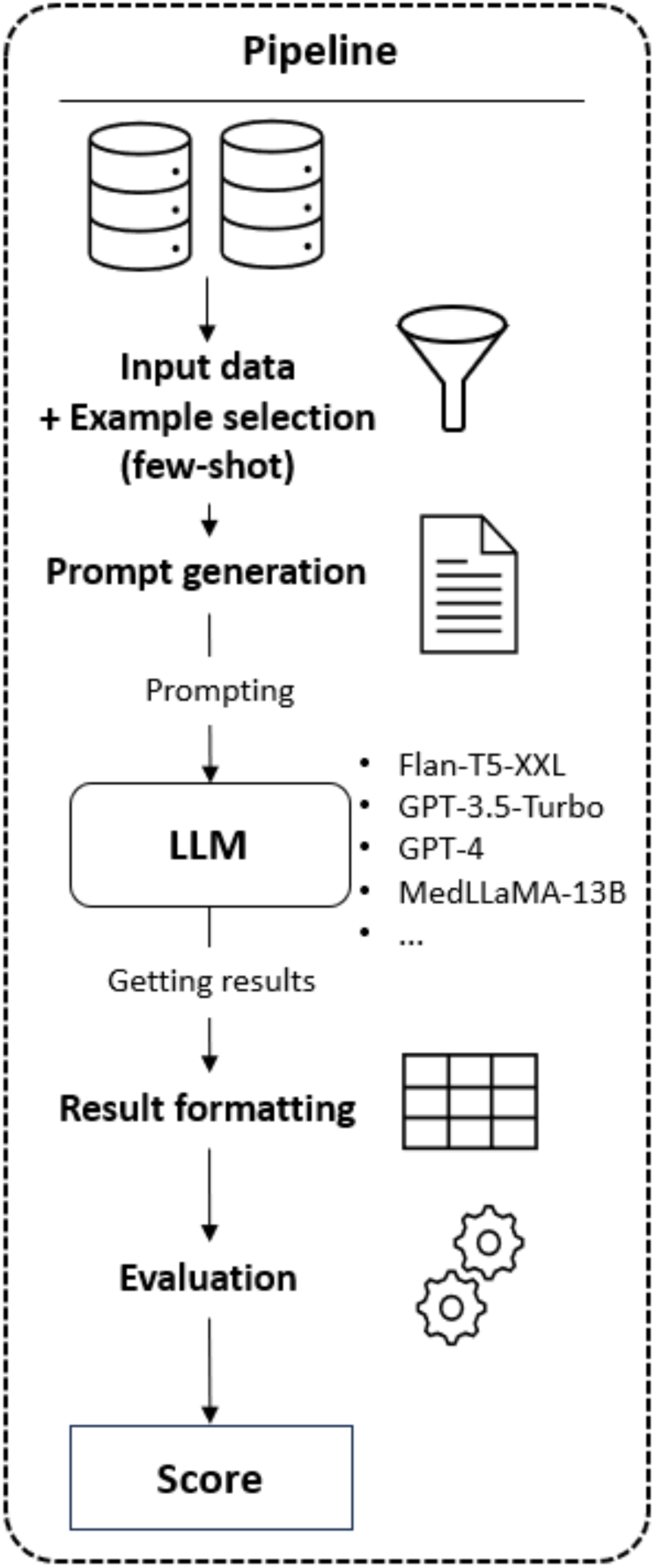
Mean performance of each large language model (LLM) for each task in the Biomedical Language Understanding and Reasoning Benchmark (BLURB) using short style versus long style zero-shot (ie, no example provided to the model) prompts (see (Multimedia Appendix 1) for more details on prompt styles and templates).

**Figure 3.**
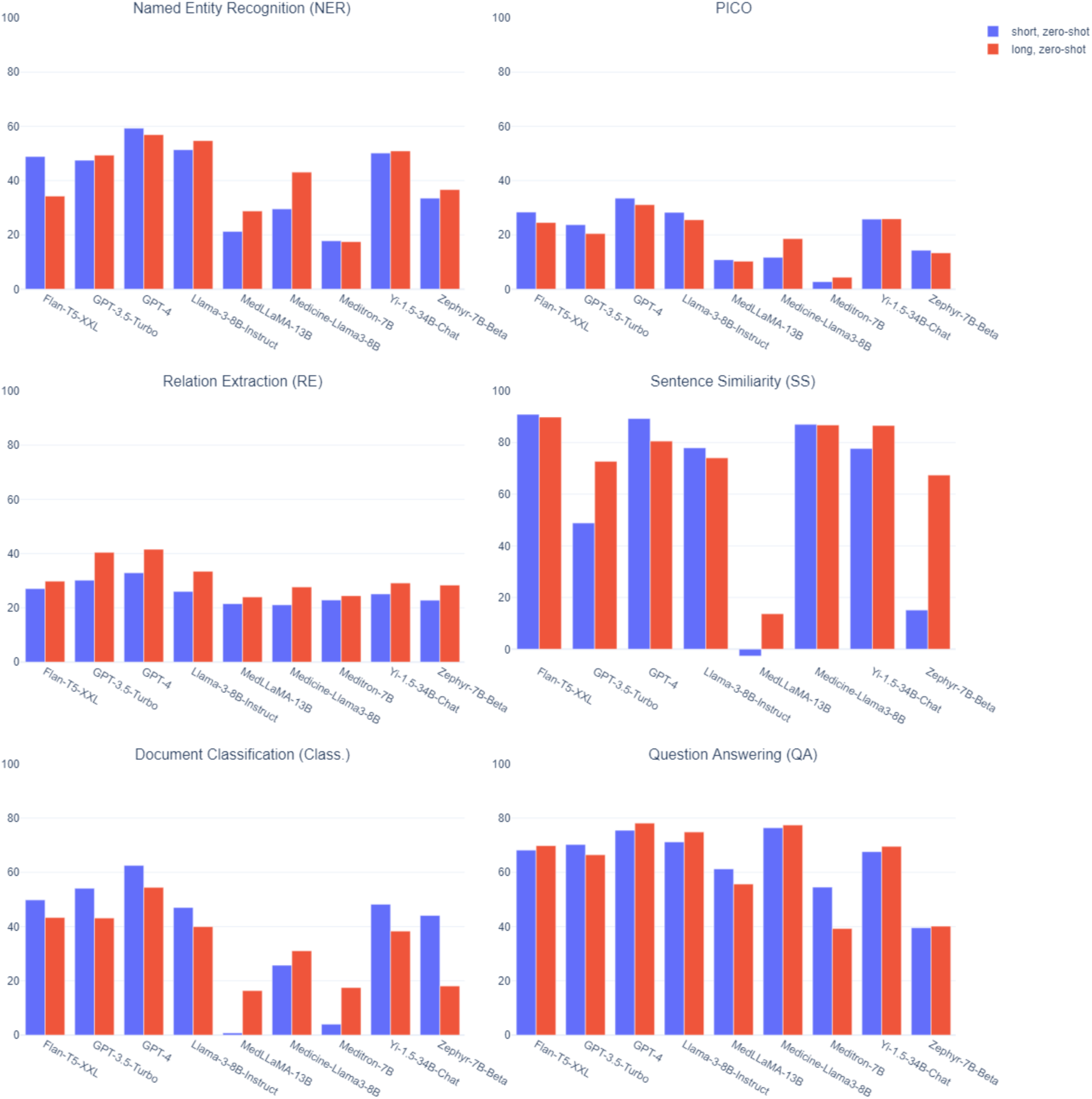
Mean performance of each large language model (LLM) for each task in the Biomedical Language Understanding and Reasoning Benchmark (BLURB) using different example selection methods: short, zero-shot: short prompt style, no example provided to the model; short, few-shot (random): short prompt style, three randomly selected examples provided to the model; short, few-shot (semantically similar): short prompt style, three examples selected based on semantic similarity provided to the model (see (Multimedia Appendix 1) for more details on prompt styles and templates).

The mean per-task performance of long versus short instructions by model when using few-shot prompting strategies is displayed in (Figure 4) (random example selection) and (Figure 5) (semantically similar example selection). Again, the effect of longer versus shorter prompts was highly dependent on the model and the task. Like zero-shot prompts, performance increased for all LLMs when using increased verbosity for the relation extraction task, regardless of example selection method.

**Figure 4.**
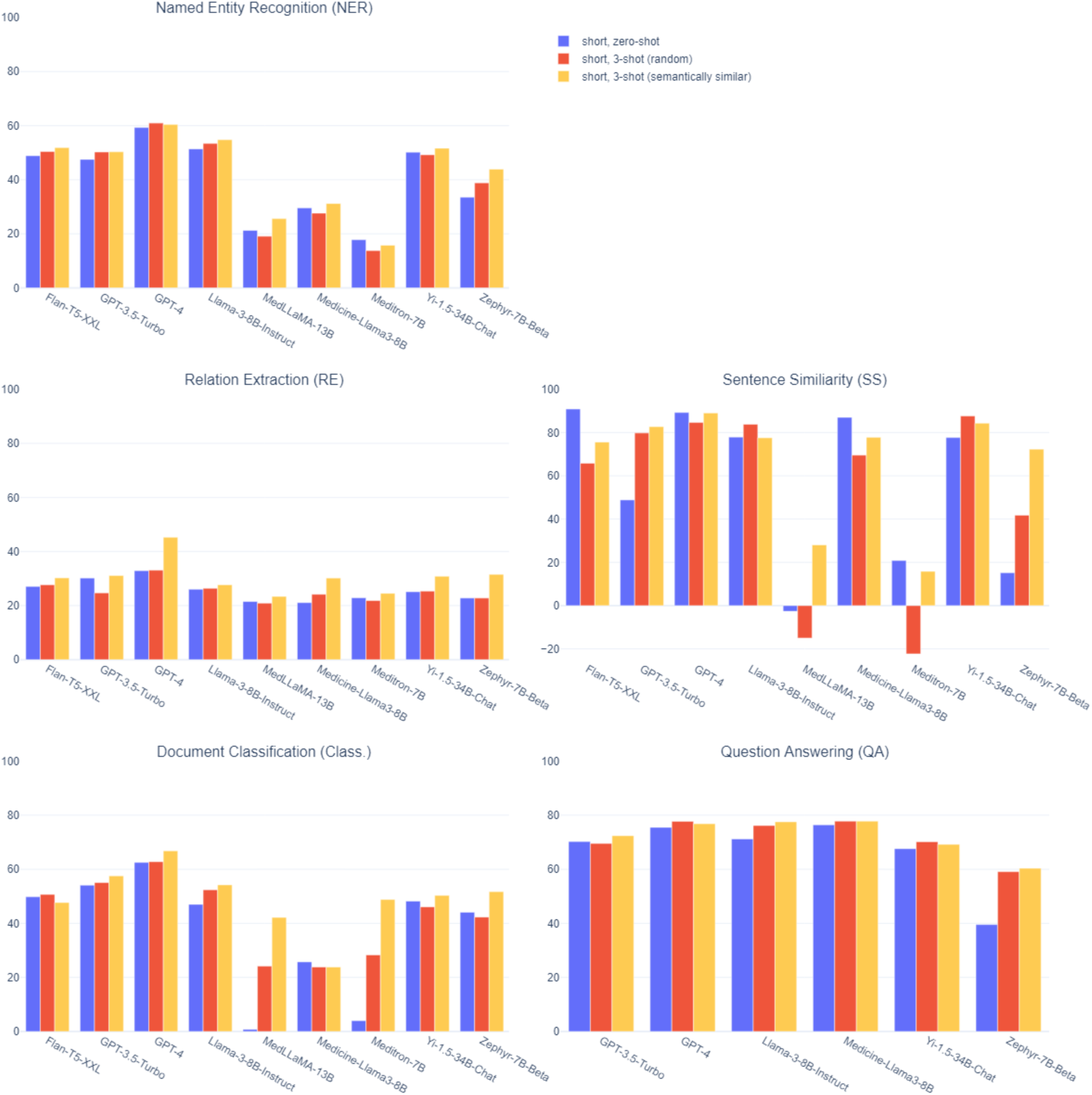
Mean performance of each large language model (LLM) for each task in the Biomedical Language Understanding and Reasoning Benchmark (BLURB) using short style versus long style random few-shot prompts (ie, three randomly selected examples provided to the model).

**Figure 5.**
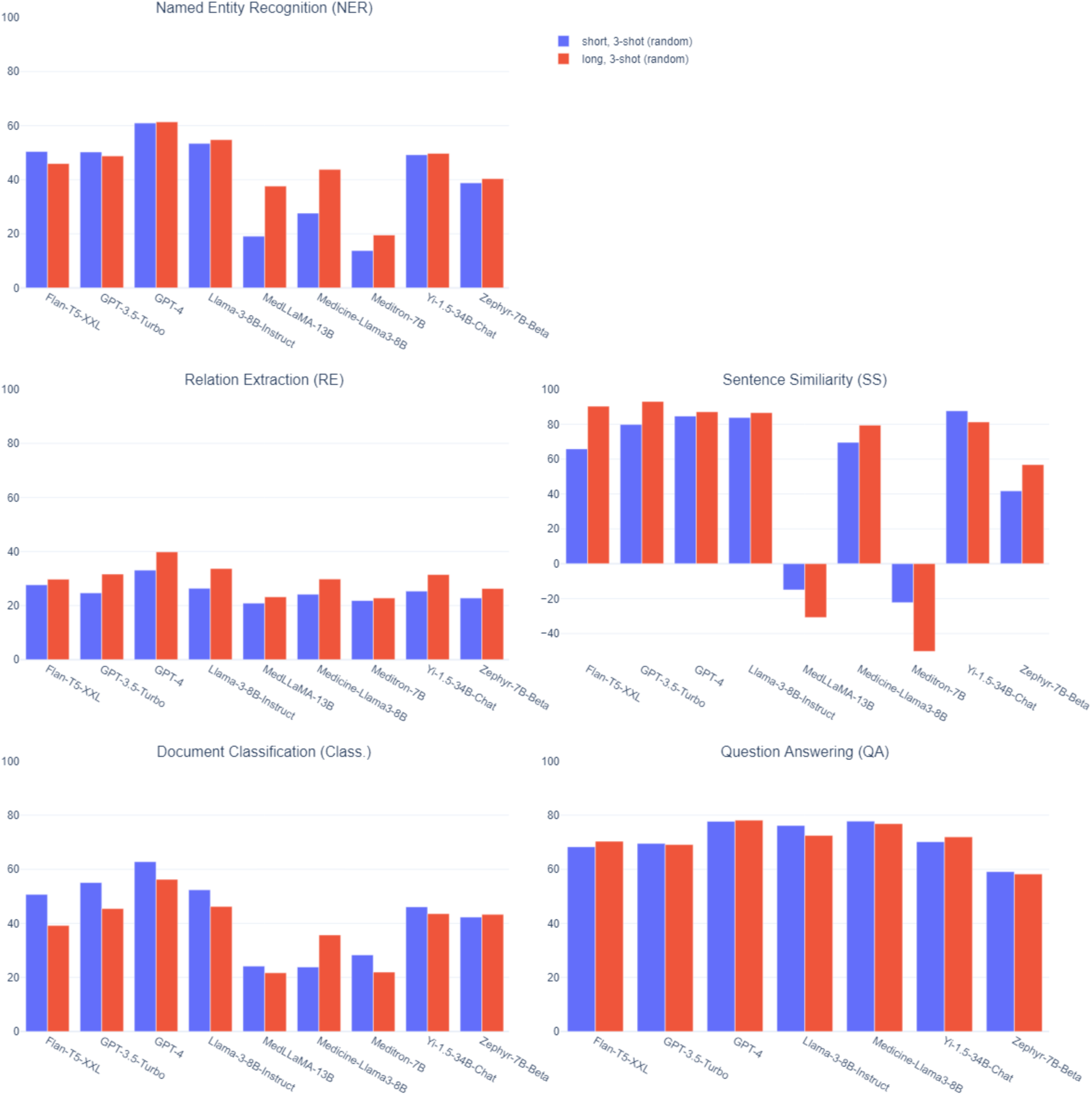
Mean performance of each large language model (LLM) for each task in the Biomedical Language Understanding and Reasoning Benchmark (BLURB) using short style versus long style semantically similar few-shot prompts (ie, three examples selected based on semantic similarity provided to the model).

### Prompting Strategies: Impact of Zero-Shot Versus Random Few-Shot Versus Semantically Similar Few-Shot

(Figure 3) compares the mean per-task performance of providing no examples (ie, zero-shot), three randomly selected examples (ie, random few-shot), and three semantically similar examples (ie, semantically similar few-shot) in the prompt when short instructions are given. For most tasks and models, the introduction of three randomly selected examples resulted in modest changes in performance compared to the zero-shot scenario. Occasionally, the differences were more substantial; for example, there were large score increases in the question answering task for Zephyr-7B-Beta and large score decreases in the sentence similarity task for MedLLaMA-13B and Meditron-7B. Overall, changes skewed slightly positive, though there were multiple tasks (NER, RE, and SS) where around half the models were negatively impacted by the addition of random examples.

Providing three semantically similar examples to the short prompt resulted in a much more consistent pattern of improvement across tasks and models. Four models (such as GPT-3.5-Turbo, MedLLaMA-13B, Yi-1.5-34B-Chat, and Zephyr-7B-Beta) experienced increased performance across all tasks, and two others (GPT-4 and Llama-3-8B-Instruct) had higher scores in all but the sentence similarity task, for which the difference was negligible. Using semantically similar examples increased the performance of every model for the RE and QA tasks, and nearly every model for NER and document classification.

Improvements ranged from modest (eg, the NER task for GPT-3.5-Turbo) to substantial (eg, the sentence similarity task for Zephyr-7B-Beta), while decreases in performance were fairly minimal outside of sentence similarity.

We observed similar patterns for long prompt scenarios; the mean per-task performance across different example selection strategies when using long prompts is compared in (Figure 6). As in short prompt scenarios, providing random examples had a modest and occasionally negative impact on the performance, while providing semantically similar examples resulted in fairly consistent improvement across all tasks and models.

**Figure 6.**
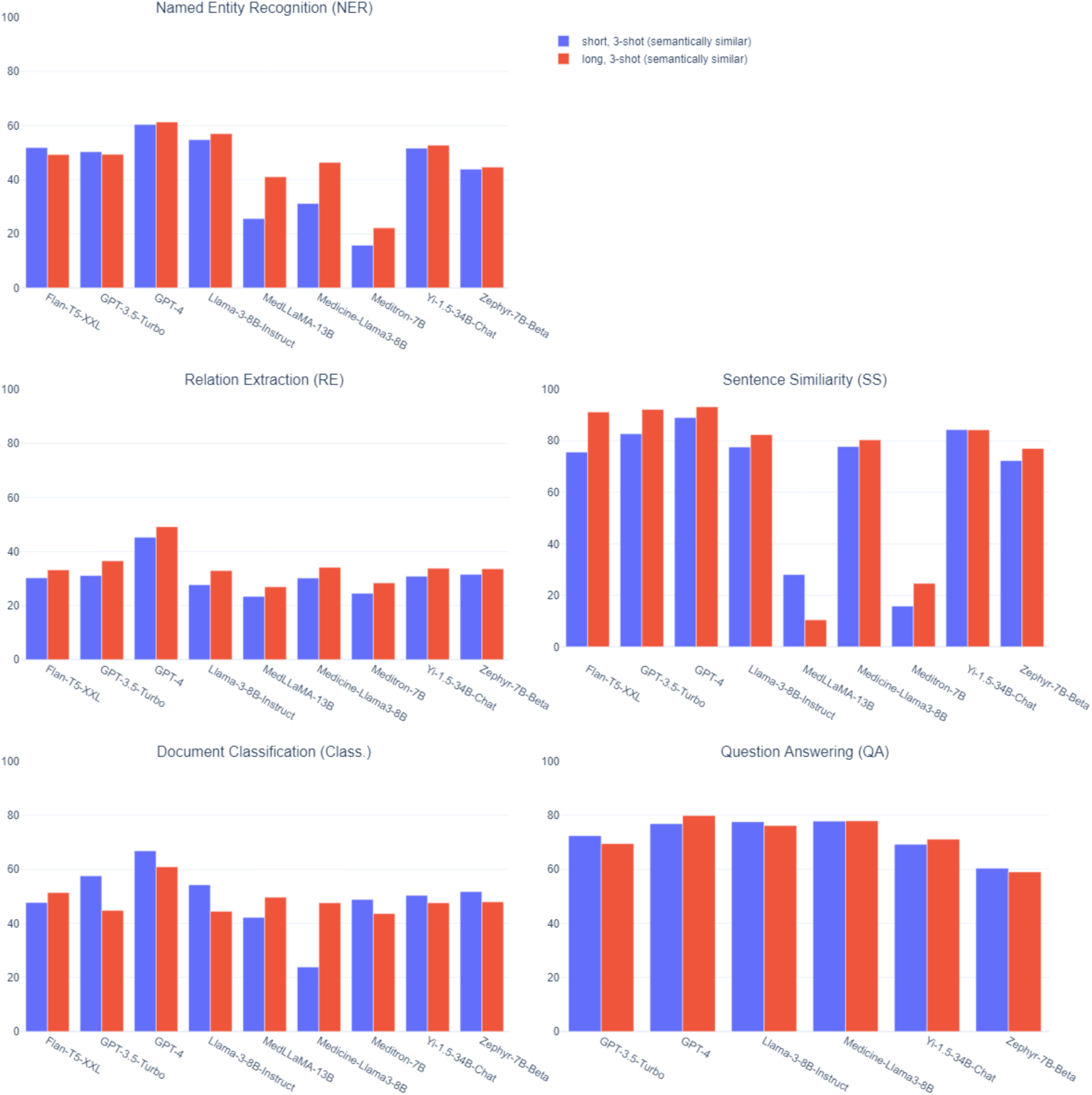
Mean performance of each large language model (LLM) for each task in the Biomedical Language Understanding and Reasoning Benchmark (BLURB) using different example selection methods: long, zero-shot: long prompt style, no example provided to the model; long, few-shot (random): long prompt style, three randomly selected examples provided to the model; long, few-shot (semantically similar): long prompt style, three examples selected based on semantic similarity provided to the model.

### Impact of LLMs

Significant differences were observed between the models’ performances. Sometimes this variation was limited to only a few tasks; for example, Llama-3-8B-Instruct scored notably better than Flan-T5-XXL and GPT-3.5-Turbo on NER and QA. In other instances, a model performed consistently better (eg, GPT-4) or worse (eg, Meditron-7B) on nearly every task.

Across 11 of the 13 BLURB datasets, GPT-4 had the highest score ((Table 4)). For all NER datasets, GPT-4 exceeded the comparators across all prompts (higher 100% of the time, with F1 scores ranging from 47.6-78.2). For BC5-chemical, BC5-disease, and NCBI-disease, the gap was large (for example, the difference between the best performing GPT-4 prompting strategy and the next best performing model for NCBI-disease was 10.01 points); for others, it was considerably smaller (for example, the difference between the best performing GPT-4 prompting strategy and the next best performing model for BC2GM was 3.73 points). For GAD, prompting Zephyr-7B-Beta with semantically similar examples performed better than any GPT-4 prompting strategy (3 points higher than the best of GPT-4). For PubMedQA, all prompting strategies for Flan-T5-XXL outperformed GPT-4, with the best Flan-T5-XXL prompt achieving a score 1.40 points higher than that of GPT-4.

**Table 4.** Top performing model-prompt combinations on each dataset of the Biomedical Language Understanding and Reasoning Benchmark (BLURB).

| Task<sup>a</sup> | Dataset | Model | Prompt | Metric | Score |
| --- | --- | --- | --- | --- | --- |
| NER | BC2GM | GPT-4 | short, 3-shot (semantically similar) | F1 entity-level | 54.70 |
|  | BC5-chemical | GPT-4 | long, 3-shot (random) |  | 78.23 |
|  | BC5-disease | GPT-4 | long, 3-shot (random) |  | 63.93 |
|  | JNLPBA | GPT-4 | long, 3-shot (semantically similar) |  | 47.55 |
|  | NCBI-disease | GPT-4 | long, 3-shot (semantically similar) |  | 70.59 |
| PICO | EBM-PICO | GPT-4 | short, zero-shot | Macro F1 word-level | 33.49 |
| RE | ChemProt | GPT-4 | long, 3-shot (semantically similar) | Micro F1 | 47.42 |
|  | DDI | GPT-4 | short, 3-shot (semantically similar) |  | 44.66 |
|  | GAD | Zephyr-7B-Beta | short, 3-shot (semantically similar) |  | 62.55 |
| SS | BIOSSES | GPT-4 | long, 3-shot (semantically similar) | Pearson coefficient | 93.18 |
| Class. | HoC | GPT-4 | short, 3-shot (semantically similar) | Average micro F1 | 66.81 |
| QA | BioASQ | GPT-4 | long, zero-shot | Accuracy | 85.71 |
|  | PubMedQA | Flan-T5-XXL | long, zero-shot |  | 76.80 |

<sup>a</sup>NER: Named-Entity Recognition; PICO: Populations, Interventions, Comparators, and Outcomes; RE: Relation Extraction; SS: Sentence Similarity; Class.: Document Classification; QA: Question Answering

Two biomedical domain models, Meditron-7B and MedLLaMA-13B, performed consistently worse than all other models, with BLURB scores of 31.42 and 36.74, respectively, well below the next best model, Zephyr-7B-Beta, which had a score of 47.26. It should be noted, however, that these LLMs were the only two not fine-tuned to follow instructions, which is likely to have had an impact. Interestingly, the third biomedical model, Medicine-Llama3-8B, generally underperformed when compared to its general-purpose equivalent, Llama-3-8B-Instruct, despite both models being derived from the same base model, Llama-3-8B. As can be seen in (Multimedia Appendix 3), for most datasets, Llama-3-8B-Instruct typically outperformed Medicine-Llama3-8B when using the equivalent prompting strategy. An exception to this is the two question-answering datasets, where the reverse is true; however, Llama-3-8B-Instruct did end up having the best scoring prompt between the two models for each of these datasets.

## Discussion

### Principal Findings

This paper evaluates multiple state-of-the-art LLMs using a standard set of benchmarking tasks and systematically compares the performance of different prompting strategies. While there is no clear ‘one-size-fits-all’ prompting strategy, our findings highlight that prompt design is a crucial aspect of the models’ performance. Our results suggest that more verbose, detailed prompts may not always be an effective strategy for improving the output of LLMs. When the additional information provided is specific to the task and is less likely routinely encountered during training, such as explaining complex relations for RE, there appears to be an appreciable impact. However, for more straightforward information, such as entity definitions for NER or how to answer questions for QA, we find the additional detail to be far less effective. While the inclusion of random examples in the prompt had a slight tendency to improve performance, it also occasionally had a significant negative impact. Overall, adding semantically similar examples to prompts proved to be the most consistent way to increase performance across tasks and models, with some variant of the semantically similar, few-shot prompt being the best performing strategy in 8 of 13 datasets.

Additionally, we find evidence suggesting that current biomedical domain LLMs may not be as suited for certain biomedical NLP tasks as state-of-the-art general-purpose models, such as GPT-4 and Llama-3-8B-Instruct. This evidence could be limited because only a small number of biomedical LLMs were evaluated, including two which were not instruction-tuned. However, while not formally evaluated in this study, we found that two other biomedical domain models that had been instruction-tuned had significant issues adhering to prompts for certain tasks; for example, PMC-LlaMA [10] answered our NER prompts as multiple-choice questions, and another model always responded in JSON. A possible explanation is that the generalizability of these models might have been reduced because of overfitting, either due to focusing only on specific tasks or on specific response formats [24] [25]. Enhancing the fine-tuning process for better performance on tasks featured in BLURB could be an interesting area to explore but is out of scope for this work.

While GPT-4 generally had the best performance on most tasks and datasets, for certain tasks, one or more smaller, open-source models performed similarly. We found that some of these models (such as Flan-T5-XXL and Llama-3-8B-Instruct) were competitive with and sometimes surpassed larger commercial models like GPT-3.5-Turbo, which typically have far more parameters.

Across tasks and models, LLMs made a sizable number of errors, indicating that they likely require some form of human oversight and correction to meet adequate quality standards for use in clinical research and medical applications. For some of the BLURB tasks, even the best LLM results had substantially lower performance than other published non-generative models, including PubMedBERT, the baseline model introduced along with BLURB benchmark [11]. GPT-4’s F1 score trailed PubMedBERT by 15.54 to 31.55 points for the named entity recognition datasets and by 24.78 to 41.46 points for relation extraction datasets. However, LLMs showed a more competitive performance on the sentence similarity and question answering datasets, especially PubMedQA. For that dataset, four models (Flan-T5-XXL, GPT-4, Llama-3-8B-Instruct, and Medicine-Llama3-8B) outperformed BioLinkBERT-Large, the leading model on the BLURB leader board [26].

### Limitations

Our study has several limitations worth noting. First, the datasets of the BLURB benchmark are primarily constructed using publicly available data from abstracts cataloged in PubMed. It is unclear how model performance on specific tasks, such as NER, would generalize to excerpts from other medical data with substantial differences in format or content. Second, heuristics (detailed in (Multimedia Appendix 2)) were used to normalize model output and enable an automated evaluation of performance. This may not account for tasks that models are able to perform in a more conversational manner, such as question-answering.

## Conclusions

In our study, we provide a comprehensive evaluation of the performance of several large language models on a variety of biomedical natural language processing tasks. We demonstrate that the optimization of prompts, namely the inclusion of semantically similar examples, can have a substantial impact on the performance of these models. Additionally, we find that, contrary to expectations, general-purpose LLMs tend to consistently outperform models specifically adapted to the biomedical domain. The heterogeneity in results across prompting strategies, models, and datasets underscores the importance of evaluating the performance of a given model and prompt for specific use cases.

Our results suggest great potential in adopting LLMs to execute biomedical tasks; notably, performant open-source models such as Llama-3-8B-Instruct show promise for cases where security, explainability, or cost are of concern. Yet our findings also show the gap between current LLMs and more traditional models for some of these tasks. Continuing to explore how to enhance the performance of these models in medical settings, paired with human expertise and quality control measures, will be important to allow responsible innovations with LLMs in the medical field.

## Supporting information

Appendix 1 - prompt template

Appendix 2 - response resolution

Appendix 3 - full table of results

## Data Availability

All underlying data used in this study are available online at https://microsoft.github.io/BLURB/tasks.html

## Acknowledgements

This study is funded by IQVIA. All authors are employees of IQVIA.

## Authors’ Contributions

HF and FR were responsible for conceptualization. JL, MG, RO, and FR contributed to the experiment methodology. HF, FR, and JN were responsible for supervision. MG, JF, RO, KRoth validated the experiment results. KRough, HF, RO, MG, JF, and FR wrote the original manuscript. JN, KZ, and CM further reviewed and edited the manuscript.

## Conflicts of Interest

FR had received research fundings from Torres Quevedo R&D Contractor, Spanish Ministry of Science, Innovation and Universities (up to 11/2021). HF, KRough, JN, CM, KZ have stock in IQVIA. RO has stock in Arria NLG. KRough has stock in Google. JN has stock in Microsoft, AZ, Nvidia, Meta. CM has stock in AZ, J&J, and MindMed. FR was previously employed by Medbioinformatics Solutions SL. RO was previously employed by Arria NLG. JN was previously employed by AZ. KRough was previously employed by Google.

## Abbreviations

BERT: Bidirectional Encoder Representations from Transformers
BLURB: Biomedical Language Understanding and Reasoning Benchmark
Class.: document classification
LLM: large language model
NER: named entity recognition
NLP: natural language processing
PICO: population, interventions, comparators, and outcomes
QA: question answering
RE: relation extraction
SS: sentence similarity

## Notes

### Competing Interest Statement

All authors are employees of IQVIA. This study is funded by IQVIA.
FR had received research fundings from Torres Quevedo R&D Contractor, Spanish Ministry of Science, Innovation and Universities (up to 11/2021). HF, KRough, JN, CM, KZ have stock in IQVIA. RO has stock in Arria NLG. KRough has stock in Google. JN has stock in Microsoft, AZ, Nvidia, Meta. CM has stock in AZ, J&J, and MindMed. FR was previously employed by Medbioinformatics Solutions SL. RO was previously employed by Arria NLG. JN was previously employed by AZ. KRough was previously employed by Google.

### Funding Statement

This study was funded by IQVIA

### Summary of Updates

We have added evaluation results on additional models, and have updated our analysis and discussions based on the additional observations.

