## Appendix 1 - prompt template for "Evaluation of Large Language Model Performance on the Biomedical Language Understanding and Reasoning Benchmark: Comparative Study"

### Multimedia Appendix: Prompt Templates

This is a Multimedia Appendix to a full manuscript published in the J Med Internet Res. For full copyright and citation information see http://dx.doi.org/10.2196/jmir.xxxx

The following are the prompt templates used for each task. Prompts differed depending on whether the LLM was a chat model:

| | **Basic** | **Chat** |
|---|---|---|
| **Models** | • Flan-T5-XXL<br>• Medicine-Llama3-8B<br>• Meditron-7B<br>• MedLLaMA-13B | • GPT-3.5-Turbo<br>• GPT-4<br>• Llama-3-8B-Instruct<br>• Yi-1.5-34B-Chat<br>• Zephyr-7B-Beta |
| **Example structure** | Example:<br>Input: <EXAMPLE-INPUT><br>Output: <EXAMPLE-OUTPUT> | **USER**<br>Input: <EXAMPLE-INPUT><br>Output:<br>**ASSISTANT**<br><EXAMPLE-OUTPUT> |

For Zephyr-7B-Beta, the system prompt was instead appended to the first user message.

#### NER

| **Prompt style** | **Prompt** |
|---|---|
| **Short, basic** | I am an excellent linguist and an expert in the biomedical domain. The task is to output a comma-delimited list of <ENTITY_TYPE> entities found in the given sentence.<br><EXAMPLES><br>Input: <INPUT><br>Output: |
| **Long, basic** | ***INPUT***<br>The input is a sentence.<br>***OUTPUT***<br>The task is to identify all <ENTITY_TYPE> mentions in the input sentence and return them in comma-separated list.<br>***DOCUMENTATION***<br><ENTITY_TYPE>: <DEFINITION><br>***EXAMPLES***<br><EXAMPLES><br>***YOUR TURN***<br>Input: <INPUT><br>Output: |
| **Short, chat** | **SYSTEM**<br>You are an expert in the biomedical domain. You are a smart named entity recognition assistant, specialized in extracting mentions of <ENTITY_TYPE> entities from text. Based on the provided sentence, answer with NO_ENTITIES if you find no mentions of <ENTITY_TYPE> entities, otherwise, answer with a |

| | |
|---|---|
| | comma-delimited list of the mentions of <ENTITY_TYPE> entities you find. Do not provide any explanation or other characters.<br>**USER**<br><EXAMPLES><br>Input: <INPUT><br>Output: |
| **Long, chat** | **SYSTEM**<br>You are an expert in the biomedical domain. You are a smart named entity recognition assistant, specialized in extracting mentions of <ENTITY_TYPE> entities from text. I will provide you the output format, the definition of the entities you need to extract, and the sentence from which to extract the entities.<br>**USER**<br>Output Format:<br>Answer with a comma-separated list of <ENTITY_TYPE > entities. If no <ENTITY_TYPE > entities are present in the sentence, answer with NO_ENTITIES. Do not provide any explanation or other characters.<br><ENTITY_TYPE>: <DEFINITION><br><EXAMPLES><br>**USER**<br>Input: <INPUT><br>Output: |

#### Entity Definitions

| Entity type | Definition |
|---|---|
| **cell** | includes cell line (a cell culture developed from a single cell and therefore consisting of cells with a uniform genetic makeup) and cell type (a classification used to identify cells that share morphological or phenotypical features) |
| **chemical** | a compound or substance that has been purified or prepared. |
| **disease** | a disorder of structure or function in a human, animal, or plant, especially one that has a known cause and a distinctive group of symptoms, signs, or anatomical changes. |
| **gene** | a basic unit of heredity and a sequence of nucleotides in DNA. |
| **protein** | a large, complex molecule that play many critical roles in a living organism. |

#### PICO

| Prompt style | Prompt |
|---|---|
| **Short, basic** | I am an excellent linguist and an expert in the biomedical domain. The task is to output a comma-delimited list of clinical trial <ENTITY_TYPE> entities found in the given clinical trial document.<br>Input: <INPUT><br>Output: |
| **Long, basic** | ***INPUT***<br>The input is a clinical trial document.<br>***OUTPUT*** |

| | |
|---|---|
| | The task is to identify all mentions of clinical trial <ENTITY_TYPE> in the input clinical trial document and return them in comma-separated list.<br>***DOCUMENTATION***<br><ENTITY_TYPE>: <DEFINITION><br>***YOUR TURN***<br>Input: <ABSTRACT><br>Output: |
| **Short, chat** | **SYSTEM**<br>You are an expert in the biomedical domain. You are a smart named entity recognition assistant, specialized in extracting mentions of outcomes of clinical trials from text. Based on the provided clinical trial report, answer with NO_ENTITIES if you find no mentions of clinical trial outcomes, otherwise, answer with a comma-delimited list of the mentions of clinical trial outcomes you find. Do not provide any explanation or other characters.<br>**USER**<br>Input: <INPUT><br>Output: |
| **Long, chat** | **SYSTEM**<br>You are an expert in the biomedical domain. You are a smart named entity recognition assistant, specialized in extracting mentions of outcomes of clinical trials from text. I will provide you the output format, the definition of the entities you need to extract, and the sentence from which to extract the entities.<br>**USER**<br>Output Format:<br>Answer with a comma-separated list of clinical trial outcomes. If no outcomes are present in the sentence, answer with NO_ENTITIES. Do not provide any explanation or other characters.<br><ENTITY>: <DEFINITION><br>Input: <INPUT><br>Output: |

#### Entity Definitions

| Entity type | Definition |
|---|---|
| **clinical trial participant** | a person who takes part in a clinical trial. |
| **clinical trial intervention** | a potential drug, medical device, activity, or procedure. |
| **clinical trial outcome** | the impact that a given intervention or exposure has on the health of clinical trial participants. |

#### ChemProt

| Prompt style | Prompt |
|---|---|
| **Short, basic** | The task is to classify relations between a chemical labeled as @CHEMICAL$ and a gene labeled as @GENE$ for a sentence. The relation must be one of 'CPR:3', 'CPR:4', 'CPR:5', 'CPR:6', 'CPR:9', or 'false'.<br><EXAMPLES><br>Input: <INPUT><br>Output: |

| | |
|---|---|
| **Long, basic** | ***INPUT***<br>The input is a sentence where the chemical is labeled as @CHEMICAL$ and the gene is labeled as @GENE$ accordingly in a sentence.<br>***OUTPUT***<br>Your task is to select one out of the six types of relations ('CPR:3', 'CPR:4', 'CPR:5', 'CPR:6', 'CPR:9', and 'false') for the gene and chemical without any explanation or other characters.<br>***DOCUMENTATION***<br><RELATION>, <DEFINITION><br>***EXAMPLES***<br><EXAMPLES><br>***YOUR TURN***<br>Input: <INPUT><br>Output: |
| **Short, chat** | **SYSTEM**<br>You are an expert in the biomedical domain. You are a smart relation extraction assistant, specialized in relation extraction between a chemical entity and a gene entity. Classify relations between a chemical labeled as @CHEMICAL$ and a gene labeled as @GENE$. The answer must be one of CPR:3, CPR:4, CPR:5, CPR:6, CPR:9, or false. Do not provide any explanation or other characters.<br>**USER**<br><EXAMPLES><br>Input: <INPUT><br>Output: |
| **Long, chat** | **SYSTEM**<br>You are an expert in the biomedical domain. You are a smart relation extraction assistant, specialized in relation extraction between a chemical entity and a gene entity. Classify relations between a chemical labeled as @CHEMICAL$ and a gene labeled as @GENE$. I will provide you the output format, the definition of class categories, and the text based on which to answer.<br>**USER**<br>Output Format:<br>Your answer must be one out of the six types of relations (CPR:3, CPR:4, CPR:5, CPR:6, CPR:9, and false) for the gene and chemical without any explanation or other characters.<br>Category Definition:<br><RELATION>, <DEFINITION><br><EXAMPLES><br>Input: <INPUT><br>Output: |

#### Relation Definitions

| Relation type | Definition |
|---|---|
| **CPR:3** | which includes UPREGULATOR, ACTIVATOR, and INDIRECT UPREGULATOR |
| **CPR:4** | which includes DOWNREGULATOR, INHIBITOR, and INDIRECT DOWNREGULATOR |
| **CPR:5** | which includes AGONIST, AGONIST ACTIVATOR, and AGONIST INHIBITOR |
| **CPR:6** | which includes ANTAGONIST |

| | |
|---|---|
| **CPR:9** | which includes SUBSTRATE, PRODUCT OF and SUBSTRATE PRODUCT OF |
| **false** | which indicates no relations |

#### DDI

| Prompt style | Prompt |
|---|---|
| **Short, basic** | The task is to classify relations between two drugs labeled as @DRUG$ for a sentence. The relation must be one of 'DDI-effect', 'DDI-mechanism', 'DDI-advise', 'DDI-false', or 'DDI-int'.<br><EXAMPLES><br>Input: <INPUT><br>Output: |
| **Long, basic** | ***INPUT***<br>The input is a sentence where the drugs are labeled as @DRUG$.<br>***OUTPUT***<br>Your task is to select one out of the five types of relations ('DDI-effect', 'DDI-mechanism', 'DDI-advise', 'DDI-false', and 'DDI-int') for the drugs without any explanation or other characters.<br>***DOCUMENTATION***<br><RELATION>: <DEFINITION><br>***EXAMPLES***<br><EXAMPLES><br>***YOUR TURN***<br>Input: <INPUT><br>Output: |
| **Short, chat** | **SYSTEM**<br>You are an expert in the biomedical domain. You are a smart relation extraction assistant, specialized in relation extraction between drug entities. Classify relations between two drugs labeled as @DRUG$. The answer must be one of DDI-effect, DDI-mechanism, DDI-advise, DDI-false, or DDI-int. Do not provide any explanation or other characters.<br>**USER**<br><EXAMPLES><br>Input: <INPUT><br>Output: |
| **Long, chat** | **SYSTEM**<br>You are an expert in the biomedical domain. You are a smart relation extraction assistant, specialized in relation extraction between drug entities. Classify relations between two drugs labeled as @DRUG$. I will provide you the output format, the definition of class categories, and the text based on which to answer.<br>**USER**<br>Output Format:<br>Your answer must be one out of the five types of relations (DDI-effect, DDI-mechanism, DDI-advise, DDI-false, and DDI-int) for the drugs without any explanation or other characters.<br>Category Definition:<br><RELATION>: <DEFINITION><br><EXAMPLES> |

| | |
|---|---|
| | Input: <INPUT><br>Output: |

#### Relation Definitions

| Relation | Definition |
|---|---|
| **DDI-mechanism** | This type is used to annotate DDIs that are described by their PK mechanism (e.g. Grepafloxacin may inhibit the metabolism of theobromine) |
| **DDI-effect** | This type is used to annotate DDIs describing an effect (e.g. In uninfected volunteers, 46% developed rash while receiving SUSTIVA and clarithromycin) or a PD mechanism (e.g. Chlorthalidone may potentiate the action of other antihypertensive drugs) |
| **DDI-advise** | This type is used when a recommendation or advice regarding a drug interaction is given (e.g. UROXATRAL should not be used in combination with other alpha-blockers) |
| **DDI-int** | This type is used when a DDI appears in the text without providing any additional information (e.g. The interaction of omeprazole and ketoconazole has been established) |
| **DDI-false** | This type is used when no DDI relation appears |

#### GAD

| Prompt style | Prompt |
|---|---|
| **Short, basic** | The task is to classify relations between a disease labeled as @DISEASE$ and a gene labeled as @GENE$ for a sentence. The response should be 1 if there is a relation or 0 if there is not.<br><EXAMPLES><br>Input: <INPUT><br>Output: |
| **Long, basic** | ***INPUT***<br>The input is a sentence where the disease is labeled as @DISEASE$ and the gene is labeled as @GENE$ accordingly in a sentence.<br>***OUTPUT***<br>Your task is to mark the relationship as either true (1) or false (0).<br>***DOCUMENTATION***<br>0: false<br>1: true<br>***EXAMPLES***<br><EXAMPLES><br>***YOUR TURN***<br>Input: <INPUT><br>Output: |
| **Short, chat** | **SYSTEM**<br>You are an expert in the biomedical domain. You are a smart relation extraction assistant, specialized in relation extraction between a disease entity and a gene entity. Classify relations between a disease labeled as @DISEASE$ and a gene labeled as @GENE$. Answer with 1 if there is a relation or 0 if there is not. Do not provide any explanation or other characters. |

| | |
|---|---|
| | **USER**<br><EXAMPLES><br>Input: <INPUT><br>Output: |
| **Long, chat** | **SYSTEM**<br>You are an expert in the biomedical domain. You are a smart relation extraction assistant, specialized in relation extraction between a disease entity and a gene entity. Classify relations between a disease labeled as @DISEASE$ and a gene labeled as @GENE$. I will provide you the output format, the definition of class categories, and the text based on which to answer.<br>**USER**<br>Output Format:<br>Answer with 1 or 0. Do not provide any explanation or other characters.<br>Category Definition:<br>0: there is no relation between the disease entity and the gene entity<br>1: there is a relation between the disease entity and the gene entity<br><EXAMPLES><br>Input: <INPUT><br>Output: |

#### BIOSSES

| Prompt style | Prompt |
|---|---|
| **Short, basic** | Provide a score from 0 to 4 signifying the similarity between two sentences.<br>Input: sentence1: <SENTENCE1> sentence2: <SENTENCE2><br><EXAMPLES><br>Input: sentence1: <SENTENCE1> sentence2: <SENTENCE2><br>Output: |
| **Long, basic** | ***INPUT***<br>The input is a pair of sentences named sentence1 and sentence2.<br>***OUTPUT***<br>The output is the semantic similarity in a continuous number from 0 (no relation) to 4 (equivalent).<br>***DOCUMENTATION***<br><SIMILARITY>, <DEFINITION><br>***EXAMPLES***<br><EXAMPLES><br>***YOUR TURN***<br>Input: sentence1: <SENTENCE1> sentence2: <SENTENCE2><br>Output: |
| **Short, chat** | **SYSTEM**<br>You are an expert in the biomedical domain. You are a smart semantic similarity scoring assistant, specialized in the scoring of semantic similarity between two sentences. Score the semantic similarity between two sentences named sentence1 and sentence2. Your score must be a number from 0 to 4 signifying the similarity between two sentences. Do not provide any explanation or other characters.<br>**USER** |

| | |
|---|---|
| | <EXAMPLES><br>Input: sentence1: <SENTENCE1> sentence2: <SENTENCE2><br>Output: |
| **Long, chat** | **SYSTEM**<br>You are an expert in the biomedical domain. You are a smart semantic similarity scoring assistant, specialized in the scoring of semantic similarity between two sentences. Score the semantic similarity between two sentences named sentence1 and sentence2. I will provide you the output format, the definition of the increments on the scoring scale, and the two sentences based on which to answer.<br>**USER**<br>Output Format:<br>Your answer is the semantic similarity expressed as a continuous number from 0 (no relation) to 4 (equivalent) without any explanation or other characters.<br>Category Definition:<br><SIMILARITY>, <DEFINITION><br><EXAMPLES><br>Input: sentence1: <SENTENCE1> sentence2: <SENTENCE2><br>Output: |

#### Similarity Definitions

| Similarity | Definition |
|---|---|
| **0** | the two sentences are on different topics. |
| **1** | the two sentences are not equivalent, but are on the same topic. |
| **2** | the two sentences are not equivalent, but share some details. |
| **3** | the two sentences are roughly equivalent, but some important information differs/missing. |
| **4** | The two sentences are completely or mostly equivalent, as they mean the same thing |

#### HoC

| Prompt style | Prompt |
|---|---|
| **Short, basic** | Provide a comma-separated list classifying the given input with zero, one, or multiple of the following hallmarks of cancer:<br>- activating invasion and metastasis<br>- avoiding immune destruction<br>- sustaining proliferative signaling<br>- resisting cell death<br>- cellular energetics<br>- genomic instability and mutation<br>- evading growth suppressors<br>- inducing angiogenesis<br>- enabling replicative immortality<br>- tumor promoting inflammation<br><EXAMPLES><br>Input: <INPUT> |

<table>
<tr><td></td><td>Output:</td></tr>
<tr><td>Long, basic</td><td>***INPUT***<br>The input is an abstract text.<br>***OUTPUT***<br>The output should be a comma-separated list, with relevant value for each class.<br>Include the class in the list if the article is related to that class.<br>Please note one article can be related to multiple classes.<br>Example output:<br>"resisting cell death","evading growth suppressors","avoiding immune destruction"<br>***DOCUMENTATION***<br>There are 10 cancer hallmarks you will need to decide whether the article is related to, including:<br><CLASS_LIST><br>***EXAMPLES***<br><EXAMPLES><br>***YOUR TURN***<br>Input: <INPUT><br>Output:</td></tr>
<tr><td>Short, chat</td><td>SYSTEM<br>You are an expert in the biomedical domain. You are a smart document classification assistant, specialized in biomedical document classification. Classify the provided abstract. If the abstract does not belong to any of the hallmarks of cancer, answer with EMPTY_LIST. Otherwise, answer with a comma-separated list consisting of one or multiple of the following hallmarks of cancer:<br><CLASS_LIST><br>USER<br><EXAMPLES><br>Abstract: <INPUT><br>Output:</td></tr>
<tr><td>Long, chat</td><td>SYSTEM<br>You are an expert in the biomedical domain. You are a smart document classification assistant, specialized in biomedical document classification. I will provide you the output format, the definition of class categories, and the abstract to classify.<br>USER<br>Output Format:<br>Answer with a comma-separated list consisting of one or multiple hallmarks of cancer.<br>Include the class in the list if the abstract is related to that class.<br>Please note one abstract can be related to multiple classes.<br>If the abstract does not belong to any of the hallmarks of cancer, answer with EMPTY_LIST<br>Category Definition:<br>There are 10 cancer hallmarks you will need to decide whether the article is related to, including:<br><CLASS_LIST><br><EXAMPLES></td></tr>
</table>

| | |
|---|---|
| | Abstract: <INPUT><br>Output: |

##### Class List

| HoC Class |
|---|
| activating invasion and metastasis |
| sustaining proliferative signaling |
| resisting cell death |
| cellular energetics |
| genomic instability and mutation |
| evading growth suppressors |
| inducing angiogenesis |
| enabling replicative immortality |
| avoiding immune destruction |
| tumor promoting inflammation |

#### BioASQ

| Prompt style | Prompt |
|---|---|
| **Short, basic** | Your task is to answer biomedical questions. Only output yes or no.<br><EXAMPLES><br>Input: <INPUT><br>Output: |
| **Long, basic** | ***INPUT***<br>The input is a question.<br>***OUTPUT***<br>Answer each question by providing one of the following options: yes, no.<br>***EXAMPLES***<br><EXAMPLES><br>***YOUR TURN***<br>Input: <INPUT><br>Output: |
| **Short, chat** | **SYSTEM**<br>You are an expert in the biomedical domain. You are a smart question answering assistant, specialized in answering biomedical questions. Only answer with yes, or no. Do not provide any explanation or other characters.<br>**USER**<br><EXAMPLES><br>Input: <INPUT><br>Output: |
| **Long, chat** | **SYSTEM**<br>You are an expert in the biomedical domain. You are a smart question answering assistant, specialized in answering biomedical questions. I will provide you the output format, and the question to answer.<br>**USER** |

| | |
|---|---|
| | Output Format:<br>Your answer must be one word: yes, or no. Do not provide any explanation or other characters.<br><EXAMPLES><br>Input: <INPUT><br>Output: |

#### PubMedQA

| Prompt style | Prompt |
|---|---|
| **Short, basic** | Your task is to answer biomedical questions using the given abstract. Only output yes, no, or maybe as answer.<br><EXAMPLES><br>Input: Question: <QUESTION> Abstract: <ABSTRACT><br>Output: |
| **Long, basic** | ***INPUT***<br>The input is a question followed by an abstract.<br>***OUTPUT***<br>Answer each question by providing one of the following options: yes, no, maybe.<br>***EXAMPLES***<br><EXAMPLES><br>***YOUR TURN***<br>Input: Question: <QUESTION> Abstract: <ABSTRACT><br>Output: |
| **Short, chat** | **SYSTEM**<br>You are an expert in the biomedical domain. You are a smart question answering assistant, specialized in answering biomedical questions. Answer based on the provided abstract. Only answer with yes, no, or maybe. Do not provide any explanation or other characters.<br>**USER**<br><EXAMPLES><br>Input: Question: <QUESTION> Abstract: <ABSTRACT><br>Output: |
| **Long, chat** | **SYSTEM**<br>You are an expert in the biomedical domain. You are a smart question answering assistant, specialized in answering biomedical questions. I will provide you the output format, the question to answer, and the abstract based on which to answer the question.<br>**USER**<br>Output Format:<br>Your answer must be one word: yes, no, or maybe. Do not provide any explanation or other characters.<br><EXAMPLES><br>Input: Question: <QUESTION> Abstract: <ABSTRACT><br>Output: |
