## Appendix 2 - response resolution for "Evaluation of Large Language Model Performance on the Biomedical Language Understanding and Reasoning Benchmark: Comparative Study"

### Multimedia Appendix: Response Resolution

This is a Multimedia Appendix to a full manuscript published in the J Med Internet Res. For full copyright and citation information see http://dx.doi.org/10.2196/jmir.xxxx

Several heuristics were used to post-process model responses so that they could be resolved to the set of possible predictions.

#### NER and PICO Tasks

1. Split the response on commas to create a list of candidate entities.
2. Tokenize each candidate entity into tokens using NLTK's *wordpunct_tokenize* function and join the tokens together with spaces.
3. Search the input text for each candidate entity.
4. If the candidate entity is found in the text, identify the start and end token of the candidate entity (partial matches are allowed at the start and end of the entity). Otherwise, ignore the candidate entity.
5. The first token of the entity not already corresponding to another entity is assigned a “B” tag; all other tokens in the entity are assigned an “I” tag.
6. All tokens not corresponding to an entity are assigned an “O” tag.

#### PICO

For each type of entity (*participant*, *intervention*, and *outcome*):

1. Split the response on commas to create a list of candidate entities.
2. Tokenize each candidate entity into tokens using NLTK's *wordpunct_tokenize* function and join the tokens together with spaces.
3. Search the input text for each candidate entity.
4. If the candidate entity is found in the text, identify the start and end token of the candidate entity (partial matches are allowed at the start and end of the entity). Otherwise, ignore the candidate entity.
5. All tokens of the entity not corresponding to another entity are assigned an “I” tag specific to that entity type (eg, “I-PAR” for *participant*).
6. All tokens not corresponding to an entity are assigned an “O” tag.

#### HoC

1. Split the response on commas to create a list of classes.
2. Convert each class to lowercase.
3. Remove all invalid classes from the response.

#### All Other Tasks

1. Obtain the first word of the response.
2. Strip any of the following characters from the start and end of the first word: *,:.”’*
3. Convert the first word to lowercase (QA tasks only).

If the word is still not a valid response, we consider the result as negative. For the PubMedQA dataset, invalid response is processed as “maybe”.
