## Appendix 3 - full table of results for "Evaluation of Large Language Model Performance on the Biomedical Language Understanding and Reasoning Benchmark: Comparative Study"

### Multimedia Appendix: Complete Table of Results

Performance of nine large language models on each of the Biomedical Language Understanding and Reasoning Benchmark (BLURB) datasets according to varying prompting strategies (best performing result appears in italics). Abbreviations: S0 – short, zero-shot; S3-R – short, 3-shot (random), S3-S – short, 3-shot (semantically similar); L0 – long, zero-shot; L3-R – long, 3-shot (random), L3-S – long, 3-shot (semantically similar).

This is a Multimedia Appendix to a full manuscript published in the J Med Internet Res. For full copyright and citation information see http://dx.doi.org/10.2196/jmir.xxxx

| Dataset | Prompt | Flan-T5-XXL | GPT-3.5-Turbo | GPT-4 | Llama-3-8B-Instruct | MedLLaMA-13B | Medicine-Llama3-8B | Meditron-7B | Yi-1.5-34B-Chat | Zephyr-7B-Beta |
|---|---|---|---|---|---|---|---|---|---|---|
| **Named entity recognition datasets (F1 score)** | | | | | | | | | | |
| BC2GM | S0 | 39.11 | 45.75 | 51.21 | 44.44 | 19.31 | 32.53 | 19.15 | 44.16 | 34.32 |
| | S3-R | 41.44 | 47.69 | 54.63 | 49.12 | 22.39 | 27.57 | 13.85 | 43.15 | 38.36 |
| | S3-S | 43.15 | 50.71 | *54.70* | 50.97 | 33.24 | 35.14 | 22.24 | 45.93 | 42.86 |
| | L0 | 34.63 | 45.56 | 50.30 | 45.04 | 29.04 | 39.27 | 18.33 | 41.48 | 32.84 |
| | L3-R | 38.17 | 46.14 | 53.18 | 48.31 | 31.03 | 37.86 | 18.05 | 42.91 | 38.54 |
| | L3-S | 41.11 | 49.21 | 53.99 | 50.58 | 41.68 | 43.65 | 27.74 | 44.95 | 43.79 |
| BC5-chemical | S0 | 65.61 | 60.64 | 75.06 | 72.21 | 27.07 | 43.43 | 23.00 | 73.02 | 49.04 |
| | S3-R | 66.98 | 66.41 | 77.35 | 75.62 | 24.55 | 31.97 | 18.66 | 69.64 | 53.16 |
| | S3-S | 65.18 | 62.55 | 76.20 | 74.50 | 27.00 | 35.97 | 16.59 | 67.53 | 57.57 |
| | L0 | 49.74 | 65.08 | 76.47 | 75.53 | 43.57 | 62.83 | 24.63 | 71.76 | 53.40 |
| | L3-R | 63.70 | 62.42 | *78.23* | 76.19 | 55.15 | 63.10 | 25.84 | 71.38 | 54.35 |
| | L3-S | 64.31 | 59.09 | 77.79 | 75.11 | 53.88 | 63.18 | 26.79 | 68.85 | 59.34 |
| BC5-disease | S0 | 52.16 | 44.61 | 60.63 | 52.32 | 21.84 | 27.60 | 16.79 | 44.99 | 30.84 |
| | S3-R | 54.05 | 48.98 | 62.15 | 53.61 | 11.23 | 21.83 | 8.77 | 48.35 | 36.03 |
| | S3-S | 54.67 | 47.00 | 56.28 | 51.26 | 13.88 | 21.49 | 7.38 | 45.84 | 37.81 |
| | L0 | 34.29 | 41.50 | 55.52 | 60.12 | 27.16 | 38.45 | 18.09 | 50.62 | 35.52 |
| | L3-R | 50.93 | 47.59 | *63.93* | 57.78 | 33.94 | 41.57 | 15.73 | 49.34 | 36.60 |
| | L3-S | 52.77 | 45.89 | 56.84 | 56.18 | 27.45 | 38.98 | 11.68 | 48.17 | 38.26 |
| JNLPBA | S0 | 35.84 | 39.12 | 44.94 | 35.39 | 15.33 | 14.82 | 13.54 | 34.33 | 22.74 |
| | S3-R | 37.99 | 40.25 | 45.43 | 36.97 | 23.85 | 32.48 | 18.44 | 37.87 | 33.13 |
| | S3-S | 40.34 | 42.01 | 45.99 | 43.26 | 27.83 | 36.13 | 13.27 | 42.99 | 38.76 |
| | L0 | 25.24 | 38.95 | 43.55 | 35.70 | 12.75 | 31.01 | 9.85 | 35.43 | 24.14 |
| | L3-R | 33.05 | 40.69 | 45.51 | 37.20 | 32.28 | 38.17 | 22.45 | 37.82 | 34.49 |
| | L3-S | 37.38 | 42.18 | *47.55* | 42.88 | 36.49 | 41.75 | 20.54 | 43.15 | 39.68 |
| NCBI-disease | S0 | 51.63 | 47.46 | 64.67 | 52.63 | 22.85 | 29.59 | 16.49 | 54.51 | 30.64 |
| | S3-R | 51.78 | 47.96 | 65.18 | 51.67 | 13.44 | 24.39 | 9.31 | 47.07 | 33.57 |
| | S3-S | 56.10 | 49.39 | 68.97 | 53.94 | 26.07 | 27.31 | 19.39 | 55.99 | 42.56 |
| | L0 | 27.58 | 55.72 | 58.95 | 57.30 | 31.41 | 43.96 | 16.68 | 55.35 | 37.51 |
| | L3-R | 44.10 | 47.19 | 65.98 | 54.47 | 35.76 | 38.24 | 15.60 | 47.28 | 37.91 |
| | L3-S | 50.87 | 50.39 | *70.59* | 60.58 | 45.88 | 44.57 | 24.08 | 58.65 | 42.14 |
| **Populations, interventions, comparators, outcomes task (macro F1 score)** | | | | | | | | | | |
| EBM-PICO | S0 | 28.42 | 23.78 | *33.49* | 28.28 | 10.80 | 11.71 | 2.75 | 25.84 | 14.32 |
| | L0 | 24.56 | 20.46 | 31.11 | 25.56 | 10.29 | 18.64 | 4.37 | 25.92 | 13.36 |
| **Relation extraction datasets (micro F1 score)** | | | | | | | | | | |
| ChemProt | S0 | 14.97 | 4.17 | 11.08 | 15.44 | 7.49 | 2.39 | 6.79 | 6.96 | 4.24 |
| | S3-R | 16.08 | 6.81 | 16.62 | 10.46 | 5.40 | 8.33 | 6.14 | 8.05 | 5.33 |
| | S3-S | 17.63 | 7.53 | 31.61 | 12.87 | 12.40 | 19.00 | 14.71 | 13.82 | 13.31 |
| | L0 | 20.33 | 31.51 | 38.25 | 26.47 | 7.91 | 16.61 | 7.60 | 22.88 | 19.22 |
| | L3-R | 19.94 | 26.46 | 37.59 | 27.48 | 8.86 | 17.07 | 8.12 | 24.34 | 13.07 |
| | L3-S | 22.39 | 21.64 | *47.42* | 25.06 | 14.27 | 23.78 | 14.38 | 24.44 | 19.57 |
| DDI | S0 | 15.19 | 35.18 | 37.70 | 16.97 | 10.27 | 13.54 | 8.65 | 17.65 | 14.91 |
| | S3-R | 16.01 | 18.26 | 27.98 | 16.84 | 8.79 | 17.39 | 10.20 | 18.17 | 16.06 |
| | S3-S | 16.90 | 34.69 | *44.66* | 16.89 | 7.91 | 19.43 | 10.99 | 21.80 | 18.85 |
| | L0 | 18.96 | 40.97 | 34.95 | 22.10 | 12.48 | 15.47 | 12.07 | 16.97 | 18.90 |
| | L3-R | 19.46 | 20.76 | 29.12 | 22.10 | 9.86 | 21.65 | 11.96 | 21.82 | 18.97 |
| | L3-S | 19.75 | 36.53 | 40.90 | 20.85 | 14.27 | 25.54 | 18.99 | 22.43 | 19.62 |
| GAD | S0 | 51.12 | 51.31 | 50.00 | 45.69 | 46.82 | 47.38 | 53.18 | 50.75 | 49.25 |
| | S3-R | 50.94 | 49.06 | 54.68 | 51.87 | 48.50 | 46.82 | 49.25 | 49.81 | 47.00 |
| | S3-S | 56.18 | 51.12 | 59.55 | 53.37 | 49.81 | 52.25 | 47.94 | 56.93 | *62.55* |
| | L0 | 50.19 | 48.88 | 51.50 | 51.87 | 51.69 | 50.94 | 53.75 | 47.75 | 47.00 |
| | L3-R | 49.81 | 47.75 | 52.81 | 51.50 | 50.94 | 50.75 | 48.31 | 48.31 | 47.00 |
| | L3-S | 57.49 | 51.50 | 59.18 | 53.00 | 52.25 | 53.18 | 51.87 | 54.49 | 61.61 |
| **Sentence similarity dataset (Pearson correlation coefficient)** | | | | | | | | | | |
| BIOSSES | S0 | 90.88 | 48.84 | 89.27 | 77.91 | -2.65 | 87.00 | 20.86 | 77.68 | 15.15 |
| | S3-R | 65.82 | 79.80 | 84.65 | 83.85 | -15.00 | 69.55 | -22.31 | 87.64 | 41.78 |
| | S3-S | 75.61 | 82.70 | 89.03 | 77.55 | 28.08 | 77.77 | 15.83 | 84.32 | 72.31 |
| | L0 | 89.86 | 72.69 | 80.53 | 74.03 | 13.71 | 86.81 | N/A [a] | 86.57 | 67.42 |
| | L3-R | 90.27 | 93.02 | 87.08 | 86.57 | -30.86 | 79.47 | -50.24 | 81.26 | 56.79 |
| | L3-S | 91.20 | 92.20 | *93.18* | 82.40 | 10.45 | 80.40 | 24.64 | 84.23 | 77.04 |

| Dataset | Prompt | Flan-T5-XXL | GPT-3.5-Turbo | GPT-4 | Llama-3-8B-Instruct | MedLLaMA-13B | Medicine-Llama3-8B | Meditron-7B | Yi-1.5-34B-Chat | Zephyr-7B-Beta |
|---|---|---|---|---|---|---|---|---|---|---|
| **Document classification dataset (micro F1 score)** | | | | | | | | | | |
| HoC | S0 | 49.81 | 54.10 | 62.52 | 47.02 | 0.79 | 25.75 | 3.99 | 48.24 | 44.11 |
| | S3-R | 50.73 | 55.09 | 62.78 | 52.40 | 24.16 | 23.83 | 28.34 | 46.10 | 42.32 |
| | S3-S | 47.69 | 57.57 | *66.81* | 54.24 | 42.18 | 23.83 | 48.82 | 50.34 | 51.74 |
| | L0 | 43.33 | 43.18 | 54.45 | 39.96 | 16.42 | 31.09 | 17.49 | 38.33 | 18.11 |
| | L3-R | 39.19 | 45.44 | 56.24 | 46.25 | 21.69 | 35.70 | 21.96 | 43.55 | 43.29 |
| | L3-S | 51.36 | 44.79 | 60.88 | 44.45 | 49.65 | 47.59 | 43.61 | 47.61 | 47.97 |
| **Question answering datasets (accuracy)** | | | | | | | | | | |
| BioASQ | S0 | 60.00 | 77.14 | 83.57 | 82.86 | 67.14 | 77.86 | 66.43 | 81.43 | 60.71 |
| | S3-R | 60.00 | 80.71 | 82.86 | 77.14 | 66.43 | 80.00 | 68.57 | 78.57 | 61.43 |
| | S3-S | 61.43 | 81.43 | 81.43 | 79.29 | 69.29 | 80.00 | 72.86 | 77.86 | 64.29 |
| | L0 | 62.86 | 70.00 | *85.71* | 79.29 | 67.14 | 79.29 | 67.14 | 83.57 | 59.29 |
| | L3-R | 64.29 | 81.43 | 82.14 | 75.00 | 67.86 | 77.86 | 67.14 | 78.57 | 57.14 |
| | L3-S | 61.43 | 78.57 | 84.29 | 77.86 | 65.71 | 80.00 | 67.86 | 76.43 | 60.00 |
| PubMedQA | S0 | 76.40 | 63.40 | 67.40 | 59.60 | 55.40 | 75.00 | 42.60 | 53.80 | 18.40 |
| | S3-R | 76.60 | 58.40 | 72.60 | 75.20 | N/A [b] | 75.60 | N/A [b] | 61.80 | 56.80 |
| | S3-S | N/A [b] | 63.40 | 72.20 | 75.80 | N/A [b] | 75.60 | N/A [b] | 60.60 | 56.40 |
| | L0 | *76.80* | 63.00 | 70.60 | 70.40 | 44.20 | 75.60 | 11.40 | 55.60 | 21.00 |
| | L3-R | 76.40 | 56.80 | 74.20 | 70.00 | N/A [b] | 75.80 | N/A [b] | 65.40 | 59.40 |
| | L3-S | N/A [b] | 60.40 | 75.40 | 74.40 | N/A [b] | 75.80 | N/A [b] | 65.80 | 58.00 |

[a] Pearson correlation could not be calculated due to all predictions being the same, leading to a zero-division error.

[b] Prompt length exceeded the maximum context size of models for the set-ups of all the few-shot experiments for EBM-PICO dataset and the few-shot experiments for PubMedQA with MedLLaMA-13B and Flan-T5-XXL; therefore we did not conduct the experiments for these set-ups, resulting in the N/A values in the table.
